# Rhythms in Longitudinal Thalamic Recordings are Linked to Seizure Risk

**DOI:** 10.1101/2025.10.03.25337281

**Authors:** Xinbing Zhang, Zachary Sanger, Thomas Lisko, Steffen Ventz, Robert A McGovern, Theoden I Netoff

## Abstract

**Objectives:** Seizure unpredictability remains a major clinical challenge for people with epilepsy. Previous works have shown that seizure risk is associated with circadian and multi-day cycles in both brain and physiological signals. However, it remains unclear whether neural activity from deep brain structures such as the anterior nucleus of the thalamus (ANT), the only FDA-approved deep-brain stimulation (DBS) target for treating medication-resistant epilepsy, exhibits similar cyclic modulation related to seizures. This study aimed to assess whether long-term local field potential (LFP) recordings from the ANT exhibit circadian and multi-day cycles that are associated with seizures that could be used to forecast seizure risk in a retrospective approach.

**Methods:** Seven participants implanted with the Medtronic Percept PC system for ANT-DBS underwent continuous at-home LFP recording of the theta/alpha (4-12 Hz) and self-reported seizure logs. Wavelet and Hilbert transforms were used to identify rhythmic cycles in LFPs. Circular statistics quantified seizure phase-locking to LFP cycles and patterns estimated from seizure diaries. Gaussian process regression (GPR) models were trained using the instantaneous phase and amplitude of these cycles to forecast short-term seizure risk.

**Results:** All participants exhibited circadian and multi-day cycles in their ANT LFPs, with seizures significantly phase-locked to some of these cycles. Seizure risk forecasting using LFP cycles achieved performance above chance (mean AUROC: 0.63 [0.57–0.69]). Incorporating the instantaneous cycle amplitude modestly improved prediction in some cases. Moreover, a substantial, though non-significant, positive correlation between circadian cycle power and seizure frequency was found in most participants, suggesting an elevated seizure risk when circadian cycles are stronger.

**Significance:** This study demonstrates that long-term LFP recordings from the ANT reflect rhythmic brain activity linked to seizure risk and may support seizure forecasting. Future studies should explore multi-modal approaches that incorporate both the phase and amplitude of cycles to improve prediction accuracy.

## 1 Introduction

Daily and multi-day periodicities in seizure occurrence among individuals with epilepsy have been well documented for centuries^1–3^. Seizures are known to cluster at specific times of day and across multi-day intervals, associated with long-term factors such as sleep^4,5^, stress^4,6^, diet^7^, and exercise^8^. These rhythmic patterns provide an opportunity to understand the mechanism underlying seizure occurrence^9–12^, and potentially improve seizure forecasting and personalized epilepsy management strategies^13–19^.

Recent advancements in long-term monitoring using implantable and wearable devices have revealed that various neural and physiological signals, including interictal epileptiform activity (IEA)^20,21^, heart rate^22^, and electrodermal activity^23^, are modulated over circadian and multi-day rhythms. Importantly, seizures often cluster at a certain phase of these cycles^20,22,23^, indicating that tracking multimodal biomarkers could improve seizure forecasting. The first in-human prospective clinical trial of seizure forecasting using long-term EEG has demonstrated the feasibility of this approach^24^.

The anterior nucleus of the thalamus (ANT) is currently the only FDA-approved deep-brain stimulation (DBS) target for treating epilepsy^25,26^. With the advent of the Medtronic Percept DBS system, simultaneous stimulation and recording of local field potential (LFP) at the implant site is now possible^27^. Using this system, we have recently observed that slow-Gamma oscillations (20–50Hz) are suppressed by high-frequency stimulation, indicating thalamic activity is modulated by the treatment^28^. A previous clinical study showed ANT-DBS modulates circadian and multi-day cycles of IEA at the seizure onset zone (SOZ)^29^. Given that seizures often cluster at certain phases of underlying biological rhythms^22,23^, monitoring thalamic neural activity may inform when a patient is at a high or low seizure risk. Fluctuations in the amplitude and phase of these cycles may indicate deviations from baseline seizure susceptibility. Therefore, understanding the dynamics of thalamic cycles could offer a potential biomarker for forecasting seizures and personalizing DBS therapy.

In this study, we tested the hypothesis that theta/alpha (4–12Hz) oscillations in ANT exhibit circadian and multi-day cycles, and that self-reported seizures occur at specific phases of these cycles and correlate with the strength of these cycles. All participants were implanted and chronically monitored using the Medtronic Percept system.

## 2 Materials and Methods

### 2.1 Subjects

Participants’ (N=7) data analyzed in this study have been collected as part of an ongoing clinical trial (#NCT05493722) approved by the University of Minnesota Institutional Review Board. All participants received the Medtronic Percept™ PC implantable pulse generator (IPG), which enables simultaneous stimulation and LFP recording. Participants were implanted with either the older Medtronic Legacy leads (Model 3389) or the latest Medtronic Sensight™ DBS leads (Model B33005) in bilateral ANTs. Participants were included if they had at-home recordings of theta and alpha (4-12Hz) Berger band lasting longer than two months and provided diaries with time-stamped seizure events.

While the clinical trial intended to evaluate the efficacy of stimulation parameter optimization, it also provided an opportunity to retrospectively analyze long-term changes in ANT LFP activity. During the trial, participants underwent multiple high-frequency (110-165 Hz) stimulation settings with varying parameters, which might contribute to the fluctuations observed in the recordings. A linear mixed effects model was used to examine potential relationships between stimulation parameters and neural activity.

### 2.2 At-Home Monitoring

The IPG estimates a Fast Fourier transform (FFT) on each 10-second LFP segment sampled at 250Hz, and the 10-minute average power of a predefined 5Hz frequency band was saved to the memory buffer^27^. Thus, the IPG records 144 LFP samples per hemisphere per day. The Medtronic Percept™ PC has a 60-day memory with a first-in-first-out buffer. Recordings were exported in JSON and downloaded at each in-clinic and at-home visit, scheduled about every 60 days.

### 2.3 Threshold Rejection and Discontinuity in Recordings

Recorded ANT LFP power was mostly around 1e3 µV^2/Hz. However, occasional overvoltage samples exceeded 1e7 µV^2/Hz and showed no correspondence with participant-reported events. As their relevance could not be verified, samples above 10^6 µV were rejected. Recordings were converted to power (dB), and a second-order autoregressive moving-average model was used to interpolate missing data. The lead contact impedances were linearly interpolated for each 30 days.

### 2.4 Seizure Diary

Seizure events were self-reported by participants using the Medtronic handheld and/or a written seizure diary. Some participants reported a single event multiple times due to unfamiliarity with the handled. To avoid repetitive event reports, seizures with an inter-seizure interval of less than 10 minutes were excluded. Additionally, we were only interested in forecasting the leading seizures of potential clusters. To identify seizure clusters, the distribution of log-transformed inter-seizure intervals was modeled using a Gaussian mixture model. A bimodal distribution is expected when seizure clusters are present, as clustered events tend to have much shorter inter-seizure intervals than others, forming a distinct peak in the histogram. A distribution was classified as bimodal if a two-component GMM had a lower Bayesian information criterion, indicating a better fit to the data, than a single-component model. The component corresponding to shorter intervals was excluded from the analysis. Additionally, seizures reported during an LFP recording gap were also excluded from the analysis. To align seizure occurrence with the 10-minute LFP recordings, each seizure time was adjusted to the nearest subsequent LFP sample time.

### 2.5 Mutual Information Permutation Test for Circadian Modulation

To evaluate whether LFP recordings exhibit significant circadian modulation, we computed the mutual information between time of day and LFPs averaged within each hour. Statistical significance was assessed using the 95th percentile of a distribution generated from 1,000 time-shuffled permutations.

### 2.6 Cycle Identification and Extraction

To identify cycles in LFP recordings, a scalogram estimated using a Morlet wavelet transform was averaged across time to obtain a power spectrum. To determine the significance of cycles, 1,000 normally distributed surrogate LFPs were generated. A local peak was considered statistically significant if its power exceeded the 95th percentile of the surrogate distribution. Circadian and multi-day cycles were rounded to the nearest integer days. For example, a cycle peak at a 5.1-day cycle would be rounded to a 5-day cycle. Cycles with a period longer than half of the longest continuous recording segment, without any gaps longer than 24 hours, were excluded from the analysis.

After identifying significant cycles, a bandpass filter was used to extract the signal at each cycle frequency with a +/− 33% window width. For example, to extract the circadian cycle (1Hz), the signal was bandpass-filtered between 0.75Hz (1.33-day) and 1.49 Hz (0.67-day). All filters used an infinite impulse response (IIR) to ensure time-causal filtering. Data were processed using MATLAB (R2024a; The MathWorks, Inc., Natick, MA). The *bandpass()* and *filter()* functions were used to design and apply an IIR filter. The *hilbert()* function was used to compute the analytic signal, from which instantaneous cycle phase and amplitude were extracted.

Additionally, circadian and multi-day patterns in the seizure diary were estimated by performing the same analysis but using simulated sinusoids with periods from 1 day to ⅕ of the recording length^13^.

### 2.7 Seizure Phase-Locking Analysis

To examine seizure timing relative to LFP cycles, we extracted the instantaneous cycle phase at the time of reported seizure events. To quantify the strength of phase-locking of seizures to LFP cycles, we used the phase-locking value (PLV) computed using the *circ_r()* function^30^. Statistical significance was assessed using the Rayleigh test via the *circ_rtest()*^30^. Only LFP cycles or seizure diary-derived patterns that showed significant phase-locking were included in subsequent seizure forecasting analyses.

### 2.8 Seizure Risk Modeling and Forecasting

A Gaussian process regression (GPR) was used to model the non-linear relationship between the seizure occurrence and the instantaneous phase and amplitude of the neural cycles. Phase and amplitude estimated using the Hilbert transform at each 10-minute timestamp were used to model seizure risk. We evaluated three GPR-based models that were trained to capture time-varying seizure probability as a function of identified cycles:

● Model 1 used only the LFP cycle phase.
● Model 2 incorporated both the LFP cycle amplitude (in dB) and phase.
● Model 3 used the phase of simulated sinusoids based on cycles found in the seizure diary without neural recordings, as described in Karoly et. al (2020)^13^.

Due to the limited number of seizure events, each model included no more than two cycles for seizure risk modeling and forecasting. If both hemispheres exhibited circadian LFP cycles phase-locked to seizures, the cycle with the higher PLV to seizures was used. Additionally, the multi-day cycle with the highest PLV was included.

For the GPR model, a Matern 3/2 kernel with automatic relevant determination was used. To ensure numeric stability during model optimization, the length scale parameters were initialized to 2 and 0.5 for modeling cycle amplitude and cycle phase, respectively. Model performance was evaluated using five-fold cross-validation with folds split chronologically to preserve the temporal structure of the recordings. Each fold uses 20% of the data for testing, regardless of the number of seizures, without random shuffling or stratification by seizure samples. Cycle amplitude and phase inputs were z-scored using the training set within each fold. Forecasting performance was quantified using the area under the receiver operating characteristic curve (AUROC) across the five folds.

### 2.9 Circadian Cycle Modulation and Seizure Frequency

To examine the longitudinal change in the circadian cycles, we estimated the monthly power of circadian cycles in each hemisphere. This was done by averaging the power of the Morlet Wavelet transform centered around the circadian frequency (+/− 33% bandwidth). To account for impedance-related variations in signal amplitude, circadian cycle power was normalized by the average impedance of the two recording contacts per hemisphere (Sensight: Left-0 & 7, Right-8 & 15; Legacy: Left-0 & 3, Right-4 & 7).

Monthly seizure frequency was calculated from self-reported seizure logs before the seizure cluster removal described in Section 2.4. Association between seizure frequency and circadian power in the dominant hemisphere (i.e. the hemisphere with stronger circadian modulation) was assessed using two approaches: (1) a linear regression model fitted across all participants, with data z-scored within participant; and (2) Pearson correlation coefficients computed within each participant’s data. To reduce noise, a 3-point moving average was applied to the circadian cycle power and monthly seizure frequency in the within-participant analyses.

To investigate potential factors contributing to changes in circadian amplitude, we fit a linear mixed-effects model with stimulation parameters (frequency, pulse width, and amplitude) and time (month) as fixed effects, and participant as a random effect (intercept). The hemisphere with the higher circadian cycle power averaged across time was selected. The monthly circadian amplitudes were z-scored within each participant.

### 2.9 Data availability

Data can be made available upon reasonable request to the corresponding author.

## 3 Results

### 3.1 ANT LFP Recordings

Seven participants, four (P1–P4) implanted with the Medtronic Sensight leads and three (P5–P7) implanted with the Legacy leads, were included in the study. A total of 7.37 years of at-home LFP recordings were analyzed. Demographic information of the population is shown in Table 1. Possible seizure clusters were found in 4 participants’ seizure diaries (Supplementary Fig. 1). Only the leading seizure of each cluster was included in the seizure forecasting analysis.

**Table 1 -.** Participant Demographics.

| Participant ID | Sex /Age | Recording Length (Days) | Reported Seizure # (after cluster removal) | Seizure Frequency /month (after cluster removal) | Seizure Types | Relevant history /Imaging Findings /Prior Procedures | Lead Type | Other Devices |
| --- | --- | --- | --- | --- | --- | --- | --- | --- |
| P1 | M 30s | 378 | 28 (17) | 2.2 (1.3) | 1. Focal impaired consciousness<br>2. Focal impaired consciousness -> tonic-clonic | Traumatic Brain Injury; Primary Left Frontal Encephalomalacia, Smaller Areas of Right Temporal & Occipital Encephalomalacia | Sensight | No |
| P2 | M 50s | 535 | 123 (123) | 6.9 (6.9) | 1. Focal preserved consciousness<br>2. Focal impaired consciousness with observable manifestations: distal and proximal automatisms<br>3. Generalized tonic-clonic | Left Inferior Temporal Gyrus Topectomy<br>Left Inferior Parietal/Occipital Topectomy | Sensight | VNS (OFF) |
| P3 | F 20s | 598 | 26 (26) | 1.3 (1.3) | 1. Focal preserved consciousness<br>2. Focal impaired consciousness | Perinatal ICH; bilateral parietoccipital encephalomalacia | Sensight | No |
| P4 | M 50s | 545 | 60 (44) | 3.3 (2.4) | 1. Focal impaired consciousness<br>2. Focal impaired consciousness with observable manifestations: hyperkinetic behavior<br>3. Focal impaired consciousness -> generalized tonic-clonic | None | Sensight | VNS (ON) |
| P5 | F 40s | 483 | 415 (319) | 25.8 (19.8) | 1. Focal preserved consciousness with observable manifestations: aphasia<br>2. Focal impaired consciousness with observable manifestations: automatisms | Right Temporal Lobectomy | Legacy | PaceMaker (ON) |
| P6 | F 30s | 130 | 64 (54) | 14.8 (12.5) | 1. Focal preserved consciousness<br>2. Focal impaired consciousness<br>3. Focal impaired consciousness -> generalized tonic-clonic | Parietooccipital encephalomalacia | Legacy | RNS (ON) |
| P7 | F 40s | 156 | 21 (21) | 4 (4) | 1. Focal preserved consciousness<br>2. Focal impaired consciousness<br>3. Focal impaired consciousness -> generalized tonic-clonic | None | Legacy | VNS (ON) |

Power fluctuations and rhythmic patterns of ANT local field potential (LFP) were observed in all participants. In P1, a characteristic pattern of increased LFP power followed by suppression was found around self-reported events, which were primarily tonic-clonic seizures (Fig. 1b). Similar patterns were observed in P4 and P5 (Supplementary Fig. 2), suggesting ANT LFP recordings may capture seizure-relevant neural activity. A 14-day sample revealed the circadian rhythm in P1 (Fig. 1c), whose right ANT LFP power increased during the night and decreased during the day. A similar, but weaker, pattern was seen in the left hemisphere (Fig. 1d). The mutual information between time and LFP power indicated significant circadian modulation in P1’s two hemispheres (p<.05, permutation test, Fig. 1e). Similarly, six out of seven participants, all except P2, showed significant circadian modulation of LFP powers in at least one hemisphere (Supplementary Table 1).

**Figure 1 -.**
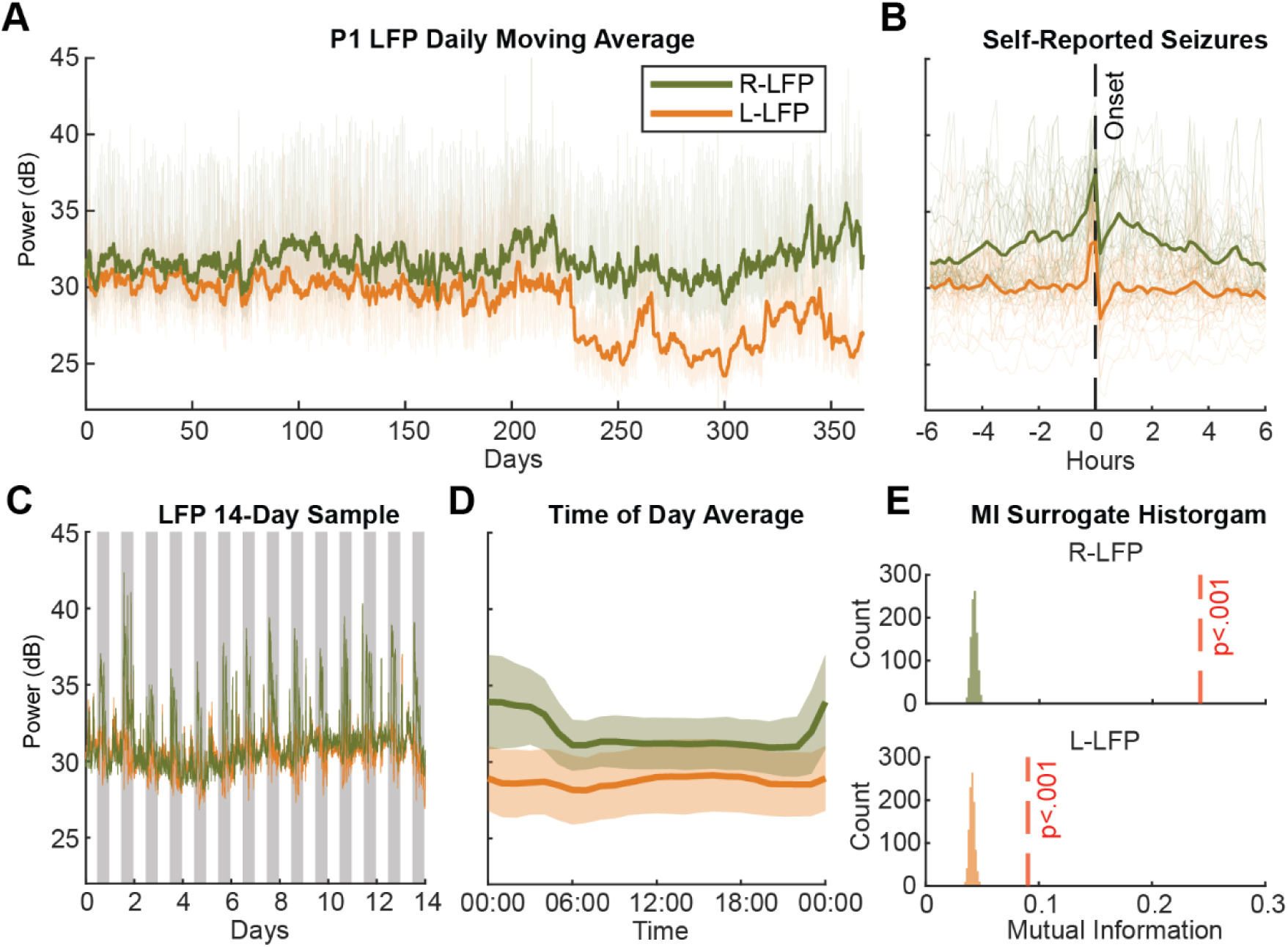
Participant 1 Example of LFP Power Recording. P1’s 10-minute average of the theta/alpha band LFP power recordings. (A) Daily moving averages of LFP powers in each hemisphere (green: right ANT; orange: left ANT). (B) 12-hour LFP recordings aligned to self-reported seizures. The x-axis shows time relative to seizure onset. (C) Recording for the first 14 days. Gray shading marks time from 8 PM to 8 AM each day. (D) LFP power averaged by time of day, highlighting consistent circadian modulation in the right ANT. (E) Histograms from the mutual information permutation test assessing circadian modulation. The red dashed line marks the mutual information using the actual data and histogram shows distribution of 1,000 permutation surrogates. Both hemispheres of P1 exhibited significant circadian modulation.

### 3.2 Circadian and Multi-day Cycles in ANT

All participants’ LFP recordings showed significant circadian and multi-day cycles (p<0.05; comparison to white-noise surrogates, Fig. 2A). Self-reported seizures were significantly phase-locked to the circadian cycle in at least one hemisphere (p<.05, Rayleigh test), indicating seizures clustered at specific phases (Fig. 2C). Although P7’s power spectrum did not exhibit a local peak at the circadian period, we found significant phase-locking of seizures to the circadian cycle in the right ANT. Across hemispheres, preferred circadian phases varied, with P1 showing marked misalignment while P3 and P5 exhibiting similar angles (Supplementary Fig. 3). Phase-locking analysis with simulated 24-h sinusoids further supported a circadian pattern in seizure timing (Figure 2D), suggesting either a diurnal or a nocturnal seizure pattern in most participants.

**Figure 2 –.**
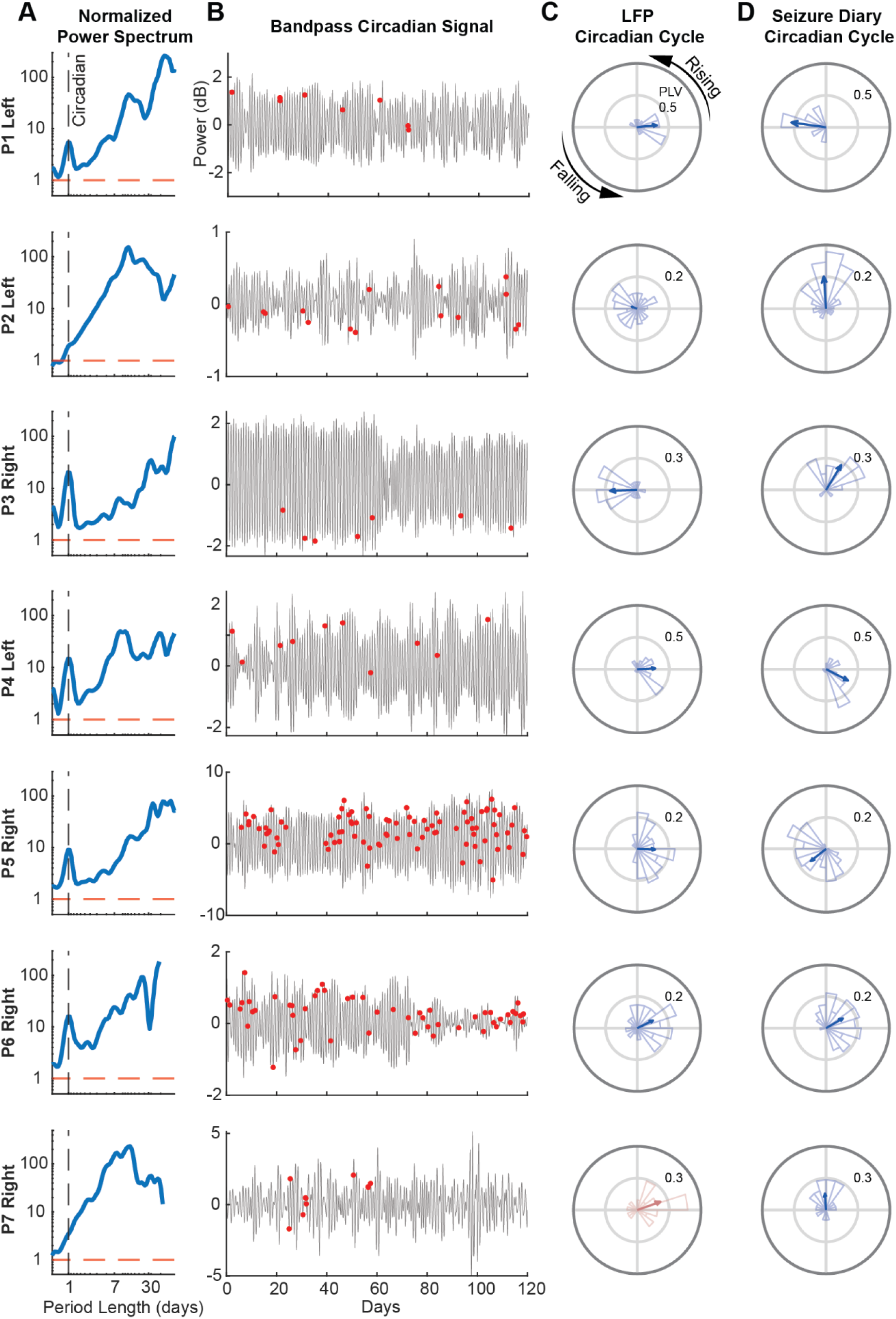
Circadian Cycle in ANT and Seizure Phase-Locking. (A) LFP scalograms averaged over time and normalized to the 95th percentile of normally distributed white-noise surrogates (red dashed line). The circadian period is marked by the vertical dashed line. For each participant example, the hemisphere with the higher phase-locking value (PLV) between its circadian cycle and seizures is shown. (B) Corresponding bandpass-filtered circadian tracings with participant-reported seizure events (red dots) for the first 120 recording days. (C) Polar histogram of seizure occurrence relative to cycle phase. The blue arrow denotes the mean resultant angle and PLV. P7 (red) did not exhibit a significant circadian cycle in either hemisphere, but seizures were phase-locked to the circadian period in the right hemisphere (p<.05, Rayleigh test). (D) Polar histograms of seizure phase-locking to a simulated 24-h sinusoid. Note that the preferred phase angle shown in (C) is estimated relative to the start of the recording. Therefore, comparisons between the two approaches should focus on amplitude (strength of phase-locking) rather than absolute phase.

For multi-day cycles, four participants had seizures significantly phase-locked to specific LFP cycles (Figure 3A). In P1 and P5, cycles of the same period length were observed across hemispheres, with seizures clustering at similar preferred phases. Some of these cycles were also captured in the seizure diary (Figure 3B), while others were identified only in the LFP recordings. This suggests that LFP- and diary-based monitoring systems capture similar but distinct seizure-related rhythms.

**Figure 3 –.**
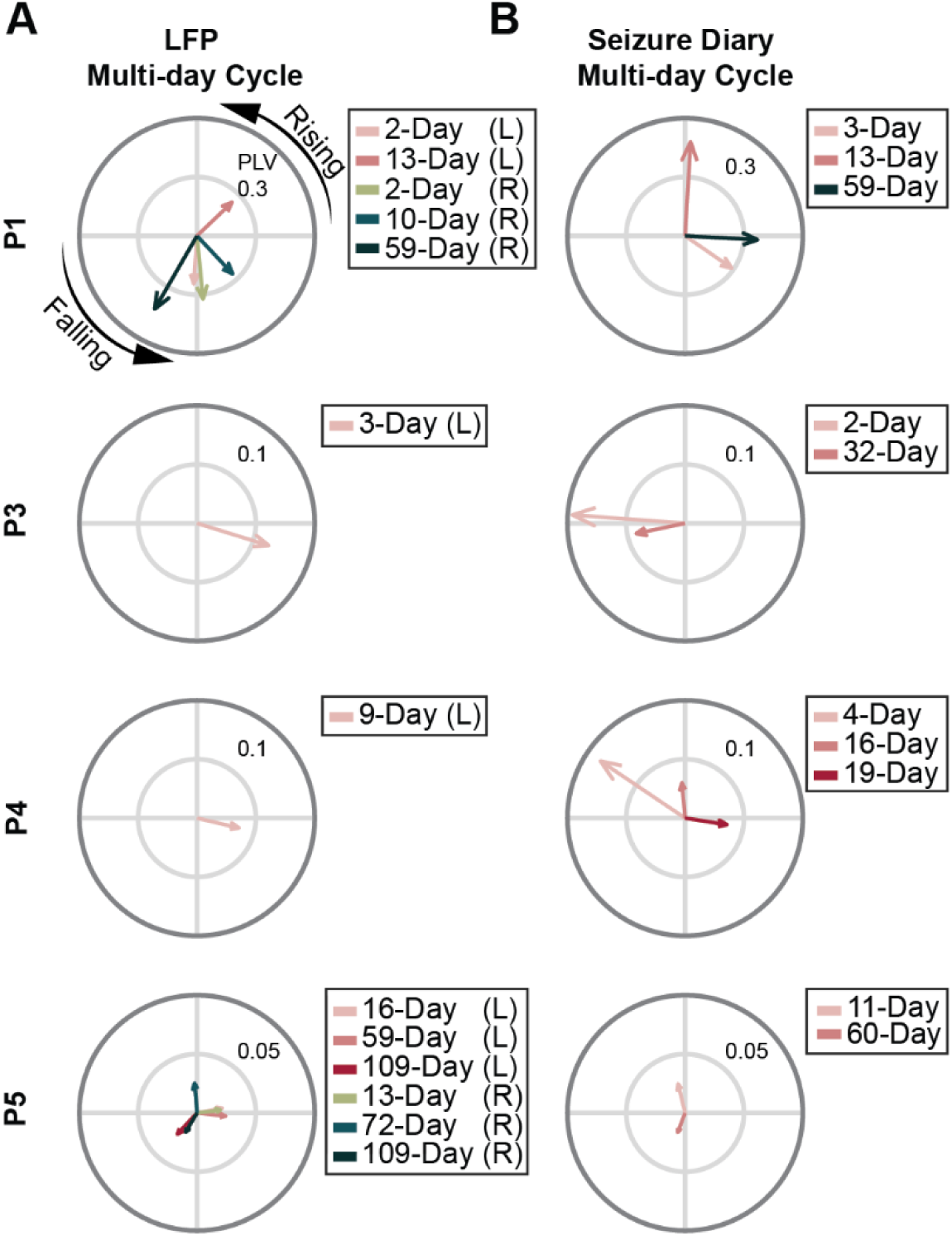
Seizures Phase Locking to Multi-day Cycles in ANT. (A) Polar plots of LFP multi-day cycles phase-locked to seizures. Recording from left and right ANTs were analyzed separately and labeled as ‘L’ and ‘R’ in the legends, respectively. Similar multi-day cycles were found across hemispheres in P1 (2-day cycle) and P7 (109-day cycle), and exhibited similar preferred phase angles. (B) Polar plots of multi-day patterns in the seizure diary. For P1 and P5, only patterns with a similar period length to the LFP cycles were shown. 13-day and 59-day in P1, and 59 / 60-day in P5 were found in both LFP recordings and their seizure diaries. However, most LFP cycles have distinct period lengths to the patterns found in the seizure diary. Once again, for all cycles, the phase is arbitrary, so only the amplitude should be compared.

### 3.3 Seizure Forecasting Performance

Seizure forecasting was performed using three Gaussian process regression (GPR) models based on: (1) the phase of LFP cycles, (2) both phase and amplitude of LFP cycles, and (3) the phase of seizure cycles derived from seizure diaries. Across participants, models based on LFP cycles achieved a forecasting performance better than chance (AUROC; Model 1: 0.56 [95% CI: 0.53–0.60], Model 2: 0.63 [95% CI: 0.57–0.69]), indicating that LFP cycles contain predictive information about seizure occurrence (Fig. 4). Incorporating the instantaneous cycle amplitude improved performance in several participants. While forecasting seizures using patterns derived from the seizure diary without neural information achieved the best performance overall (Model 3: 0.65 [95% CI: 0.59–0.71]), the advantage over the LFP-based model using both phase and amplitude was marginal.

**Figure 4 -.**
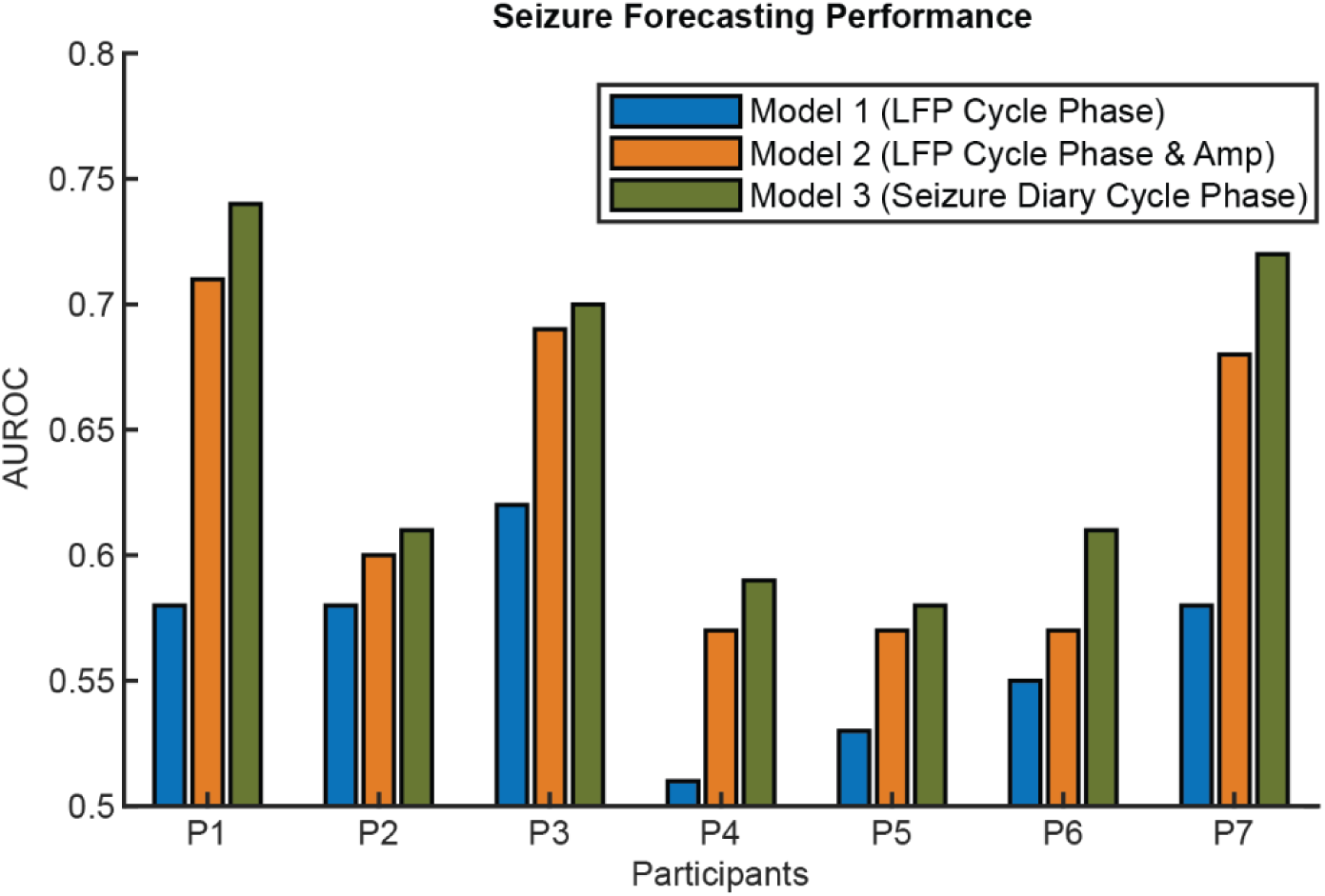
Seizure Forecasting AUROC. Results of 3 GPR-based seizure forecasting methods. Model 1: using only the phase of LFP cycles. Model 2: using both phase and amplitude of LFP cycles to forecast seizure risk. Model 3: using seizure diary-based seizure phase distribution with respect to periodic circadian and multi-day cycles. P7 did not have any LFP cycles exhibited as a local peak on their LFP power spectrum that were also phase-locked to seizures. However, the circadian cycle in their right ANT, which was not captured on the power spectrum, was phase-locked to seizures and used for forecasting here.

### 3.4 Circadian Cycle Power and Seizure Frequency

Fluctuations in circadian cycle power were found in most patients (Fig. 5a). To explore whether stimulation parameters influenced circadian modulation in the ANT, a linear mixed-effects model was fitted. The results showed a significant relationship (p<.05) between the stimulation pulse width (µs) and circadian cycle power in the dominant hemisphere (Supplementary Table 4), indicating that shorter pulse widths were linked to reduced circadian modulation in the ANT (□=0.01).

**Figure 5 -.**
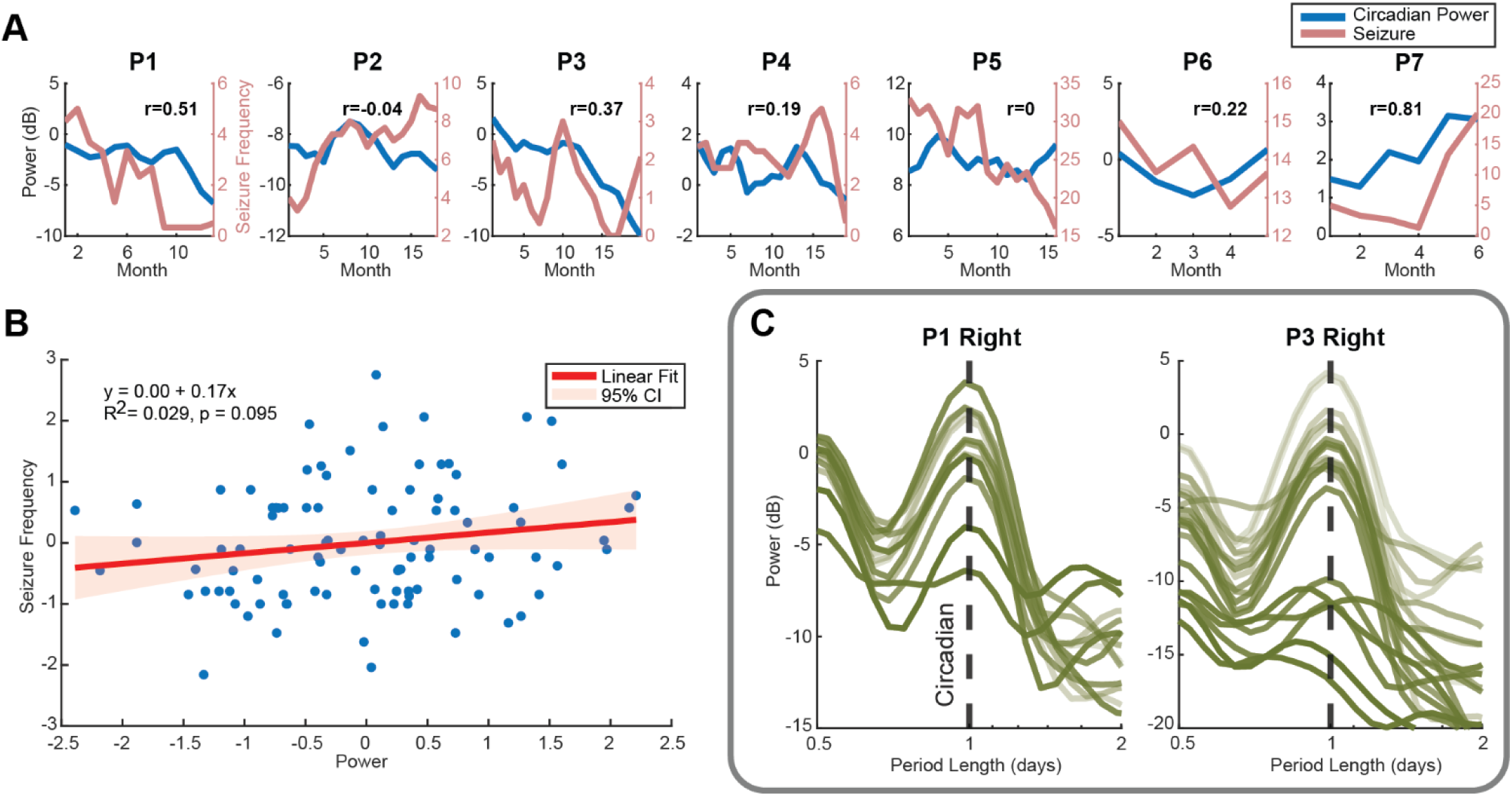
Monthly Circadian Cycle Power and Seizure Frequency. (A) 3-month moving average of seizure frequency (blue) and LFP circadian cycle power in the hemisphere with higher power averaged over time. The x-axis shows time since the start of at-home monitoring of each participant. (B) A linear fit to all participants’ monthly seizure frequency and circadian cycle power shows a weak but positive correlation. (C) Monthly power spectrum of P1 and P3 right ANT LFP. Lighter lines indicate LFP recorded earlier in the study, and darker lines correspond to later recordings. In both participants, circadian cycle power decreased over time. Despite an impedance normalization having been applied to the analysis, a baseline shift may still be present.

A correlation analysis between the cycle power in the cycle-dominant hemisphere and seizure frequency revealed a positive correlation in 5 out of 7 participants, with several showing moderate correlation coefficients (R > 0.3). A positive relationship between the circadian cycle power and seizure frequency was further confirmed by a linear regression model fitted to all participants’ data (Fig. 5b), although it did not reach statistical significance (p = 0.1). Additionally, a clear reduction of the circadian amplitude was observed in P1 and P3 (Fig. 5c).

Recordings from P2 and P5 did not show a correlation between circadian amplitude and seizure frequency (Supplementary Table 2). Interestingly, P2 lacked significant non-uniform circadian modulation in the MI permutation test, while P5 exhibited the opposite circadian pattern compared to other participants (Supplementary Fig. 5). Strong cardiac artifacts present in P5’s in-clinic recordings, sampled at 250Hz, may have contributed to this atypical circadian rhythm.

## 4 Discussion

We found strong circadian and multi-day cycles in ANT LFP recordings among medication-resistant epilepsy participants. Self-reported seizures were found to cluster at specific phases of these cycles. Most participants had seizures clustered at a similar phase of the LFP circadian cycles across hemispheres, suggesting cycles across sides of ANTs might be modulated by the common underlying physiological or pathological mechanisms. While the seizure diary showed cyclic patterns similar to LFP power recordings, certain multi-day cycles in LFP were not reflected by the diary. Seizure forecasting models based on LFP cycles significantly outperformed chance in all participants. And in some participants, utilizing the instantaneous amplitude of LFP cycles improved forecasting performance. While the result did not reach statistical significance, a positive correlation was found between the monthly power of the LFP circadian cycle and seizure frequency in most participants. This is consistent with a prior RNS study that demonstrated the modulation of the circadian cycle in IEA correlated with responder outcome^29^.

Theta/alpha activity in ANT has been reported to be relevant to epilepsy and DBS outcome. Hupalo et al. (2018) identified 5–7 Hz theta activity in the ANT of epilepsy patients^31^, while Scherer et al. (2020) showed that ANT stimulation reduces theta-band power in scalp EEG^32^. Additionally, Chaitanya et al. (2020) reported phase-amplitude coupling between low-frequency rhythms in the ANT and high-gamma activity in the SOZ^33^. Our recent case study also demonstrated seizure reduction associated with suppressed ANT theta/alpha^34^. Findings here suggest that not only is seizure timing linked to cycles in ANT theta/alpha activity, but also the modulation of the circadian cycle is associated with seizure frequency. While ANT is involved in the circuit of Papez, a brain network that is commonly involved in epilepsy, short-term power changes in ANT around seizure events were not consistently found among all participants. This is consistent with findings from previous studies suggesting that not all seizure networks significantly involve ANT^35^, with different epilepsy types involving different brain regions^36^.

While models based on seizure diary–derived patterns achieved forecasting performance comparable to those using LFP cycles, each approach relied on distinct multi-day cycles for prediction. This indicates that LFP and diary-based cycles may capture complementary aspects of seizure risk^37^. Additionally, all GPR models were initialized with the same kernel length-scale, which may have contributed to the similar performance observed here. Future studies incorporating other modalities may help determine whether these cycles in neural and physiological recordings contribute independently to seizure risk or simply reflect a shared underlying structure captured in seizure diary patterns.

Previous studies have shown that seizures are often phase-locked to cycles in neural and physiological signals, raising the possibility that these rhythms may modulate seizure timing^20,22,23^. To investigate this further, we examined whether the instantaneous LFP cycle amplitude, a quantitative measurement of the cycle strength, was related to seizure timing. We incorporated the instantaneous cycle amplitude into GPR models and observed enhanced performance across participants, with a few showing a notable increase in AUROC, supporting the potential utility of cycle amplitude in forecasting models. Although the result did not reach statistical significance, a substantial positive correlation between the circadian cycle power and seizure frequency was found in most participants. This suggests that the modulation of ANT cycles may indicate longer-term changes in seizure susceptibility.

The circadian rhythms in epilepsy were often linked to the sleep and awake states^38–41^. Studies have found that poor sleep quality is associated with increased seizure likelihood^4,5^, and ANT-DBS has been reported to influence sleep^42–44^, which might cause the change in the circadian cycle observed here. However, in some of the participants showing circadian power modulation, we observed that the change is more likely driven by fluctuations in daytime power, rather than a change in the typical rise of theta/alpha power at night (Supplementary Fig. 5). This suggests factors beyond sleep may also influence the circadian modulation observed in the ANT.

### 4.1 Limitations and Future Directions

Although this study uses years-long LFP power recordings, the main limitation is the limited number of participants, which may constrain its generalizability to the population. This is a retrospective study in which phase-locking analyses were performed using the entire recording period. Prospective studies identifying long-term cycles phase-locked to seizures may require a long calibration period^14,24,37,45^, and such phase-locking may change over time due to factors such as medication adjustments^46^. Future studies with enough seizure samples should consider using a pseudo-prospective approach to evaluate potential seizure biomarkers^13,37^.

Another limitation is the unreliability of seizure diaries^4,47–50^. However, a recent study found that seizure diaries can be used to estimate the underlying cycles and achieved a similar result to using EEG^51^. Nevertheless, future studies should consider utilizing wearables with the ability to detect seizures to provide better seizure tracking.

Our forecasting models did not utilize historical information. A recent study investigated a hybrid model using cycles for long-horizon forecasting achieved promising performance^37^. A model that includes historic ANT cycle information may improve the forecasting. Additionally, fixed kernel parameter initialization was applied across the three tested models to ensure convergence and numerical stability. A well-designed parameter optimization for the GPR-based models may enhance the model accuracy.

## 5 Conclusion

Our findings provide evidence that LFP cycles in the ANT are commonly observed in medication-resistant epilepsy patients, with self-reported seizures clustering at specific phases of these cycles. Incorporating the instantaneous amplitude of these cycles improved seizure forecasting performance in this retrospective analysis. Most participants exhibited a substantial association between circadian cycle modulation and clinical outcome, highlighting the potential clinical relevance of neural cycle dynamics, although this effect did not reach statistical significance in this study. Future studies with a larger cohort and a prospective approach may further clarify this association and guide the integration of cycle-based biomarkers into personalized clinical management of epilepsy.

## Data Availability

All data produced in the present study are available upon reasonable request to the authors

## Acknowledgements

Thank you to the participants and their neurologists for their contributions and participation within the study. Thank you to Scott Stanslaski for technical assistance when using the Medtronic Percept™ system within our ongoing clinical trial.

## Supplemental Materials

**Supplementary Figure 1 -.**
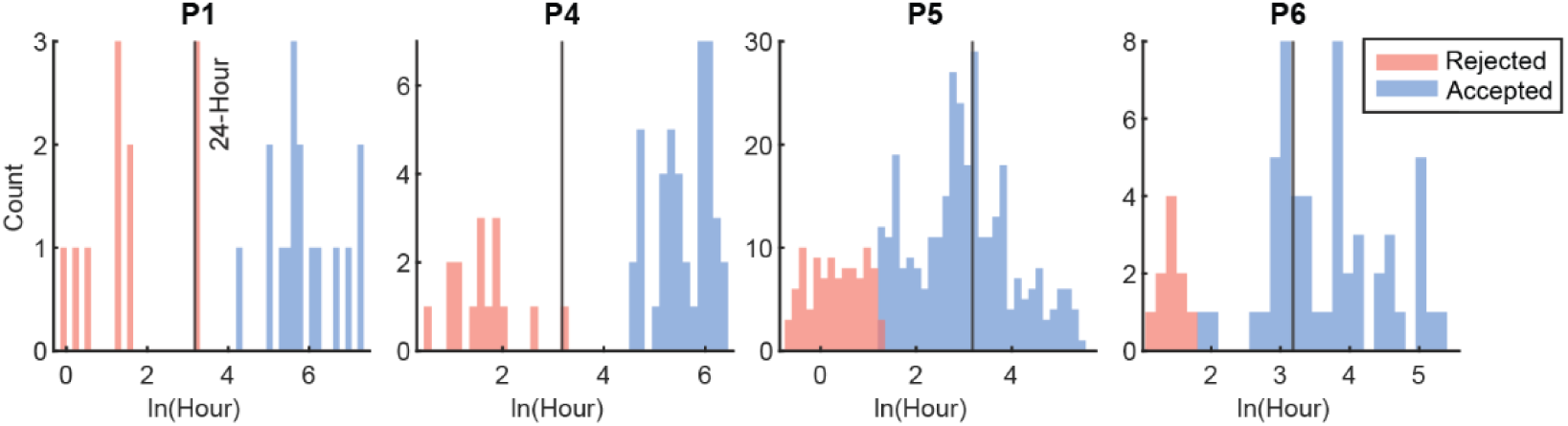
Seizure Clusters Identification. Seizures corresponding to the distribution with a shorter inter-seizure interval mean (red) were removed from the forecasting analysis. Possible seizure clusters were found in 4 participants. The x-axis shows the log-transformed inter-seizure interval. All rejected seizure events have inter-seizure intervals shorter than 24 hours.

**Supplementary Table 1 -.** Circadian LFP Rhythm Mutual Information Permutation Test. Mutual information between the hourly mean LFP power recordings and time was compared against 1,000 random permutations of the recordings. All participants except P2 exhibited significance, indicating non-uniform circadian modulation in ANT powers.

|  | Left p-value | Right p-value |
| --- | --- | --- |
| P1 | <.001 | <.001 |
| P2 | 0.7 | 0.44 |
| P3 | <.001 | <.001 |
| P4 | <.001 | <.001 |
| P5 | <.001 | <.001 |
| P6 | <.001 | <.001 |
| P7 | <.001 | 0.85 |

**Supplementary Figure 2 -.**
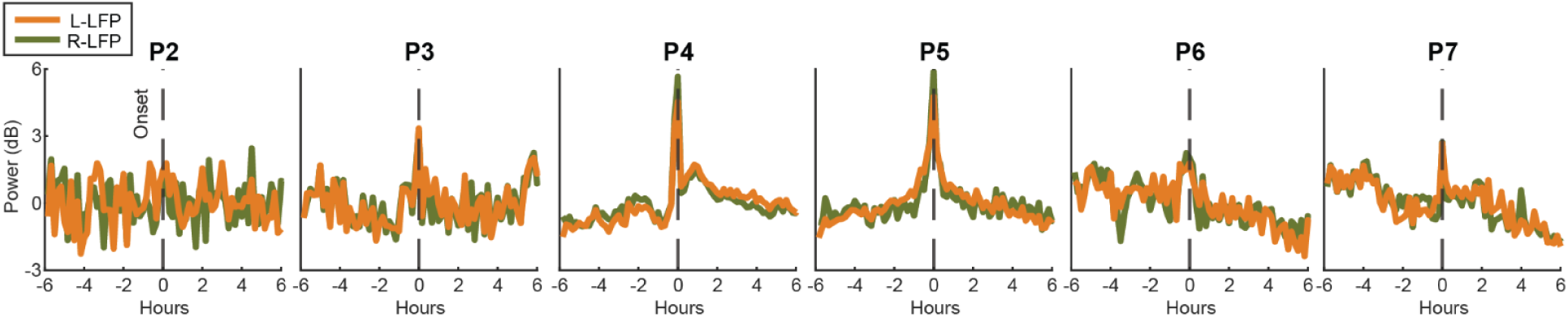
LFP Around Self-Reported Seizures. Increased theta/alpha power was found around self-reported seizures in most participants. However, the power suppression following seizures observed in P1 was not seen in other participants’ recordings.

**Supplementary Figure 3 -.**
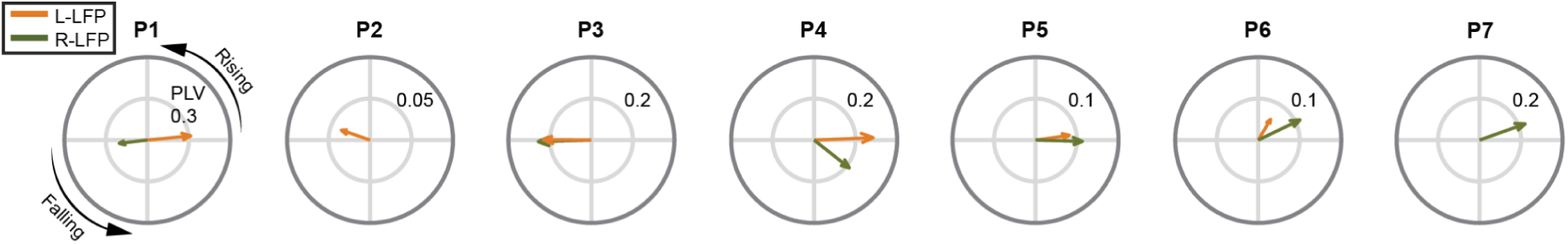
Circadian Cycles Mean Resultant Vector of Both Hemispheres. Overall seizure prevalence relative to the LFP circadian cycle phase in each hemisphere. The green and orange arrows denote the mean resultant angle and phase locking value of left and right ANT recordings, respectively. All participants, except P2 and P7, exhibited significant phase-locking in both hemispheres. Most participants had seizures clustered at a similar phase across hemispheres, but P1 exhibited opposite phase preferences between hemispheres.

**Supplementary Figure 4 -.**
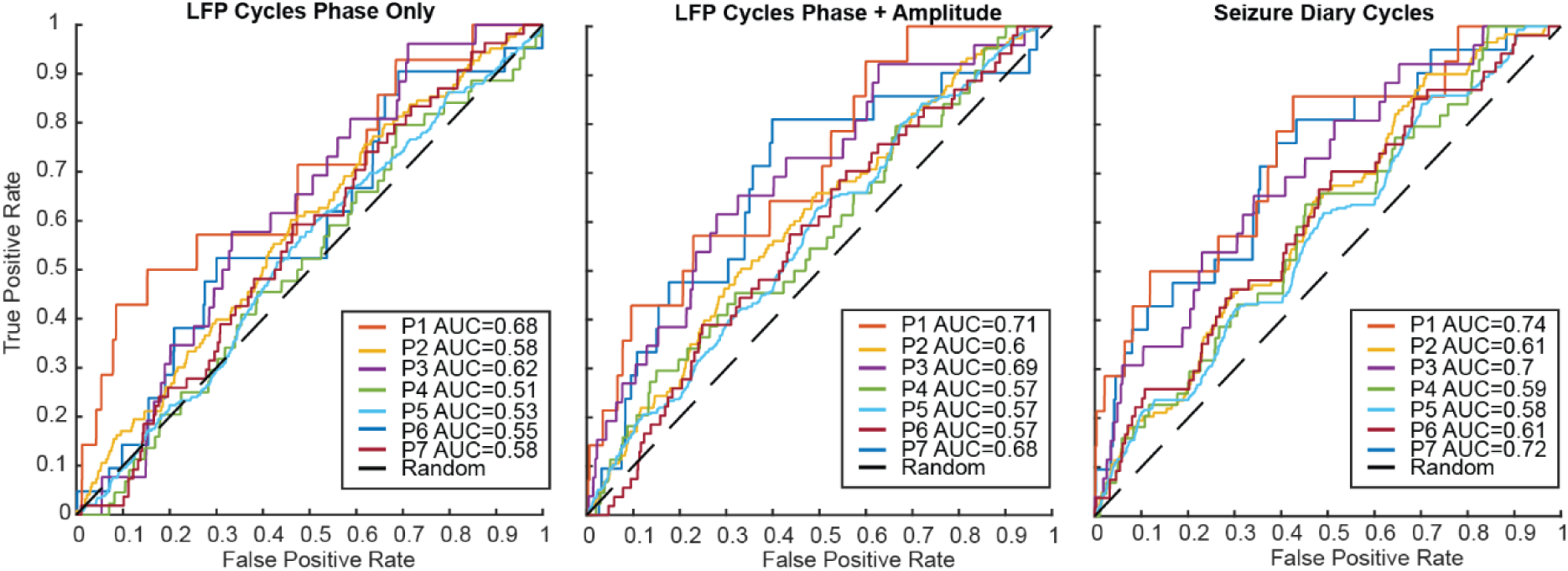
Seizure Forecasting ROCs.

**Supplementary Figure 5 -.**
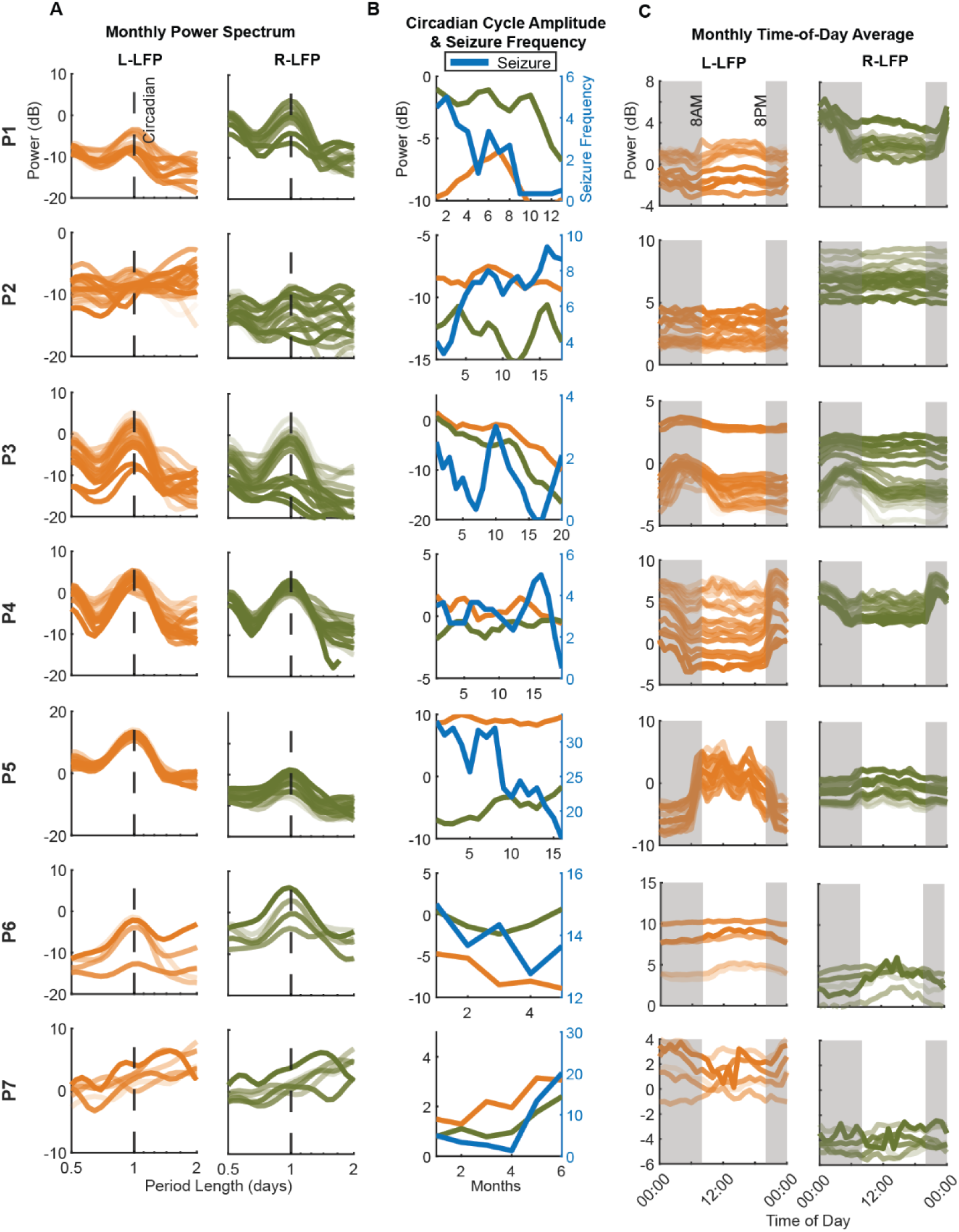
Circadian Cycle Modulation and Seizure Frequency. (A) Monthly scalograms of left and right ANT LFP powers averaged over time. Results were normalized by the impedance of recording contacts. Lines with a more transparent color indicate LFP collected earlier in the study, and vice versa. (B) 3-month moving average of circadian cycle amplitude modulation in left (orange) and right (green) hemispheres, along with seizure frequency (blue). The x-axis shows time since the start of at-home monitoring of each participant. (C) LFP averaged by time of day in each month. The same color code in (A) applies here. P1, P3, and P4 demonstrated the circadian pattern with increased activity during night (shaded in grey). Despite an impedance normalization having been applied, a potential baseline shift still exists in some participants’ results.

**Supplementary Table 2 -.**
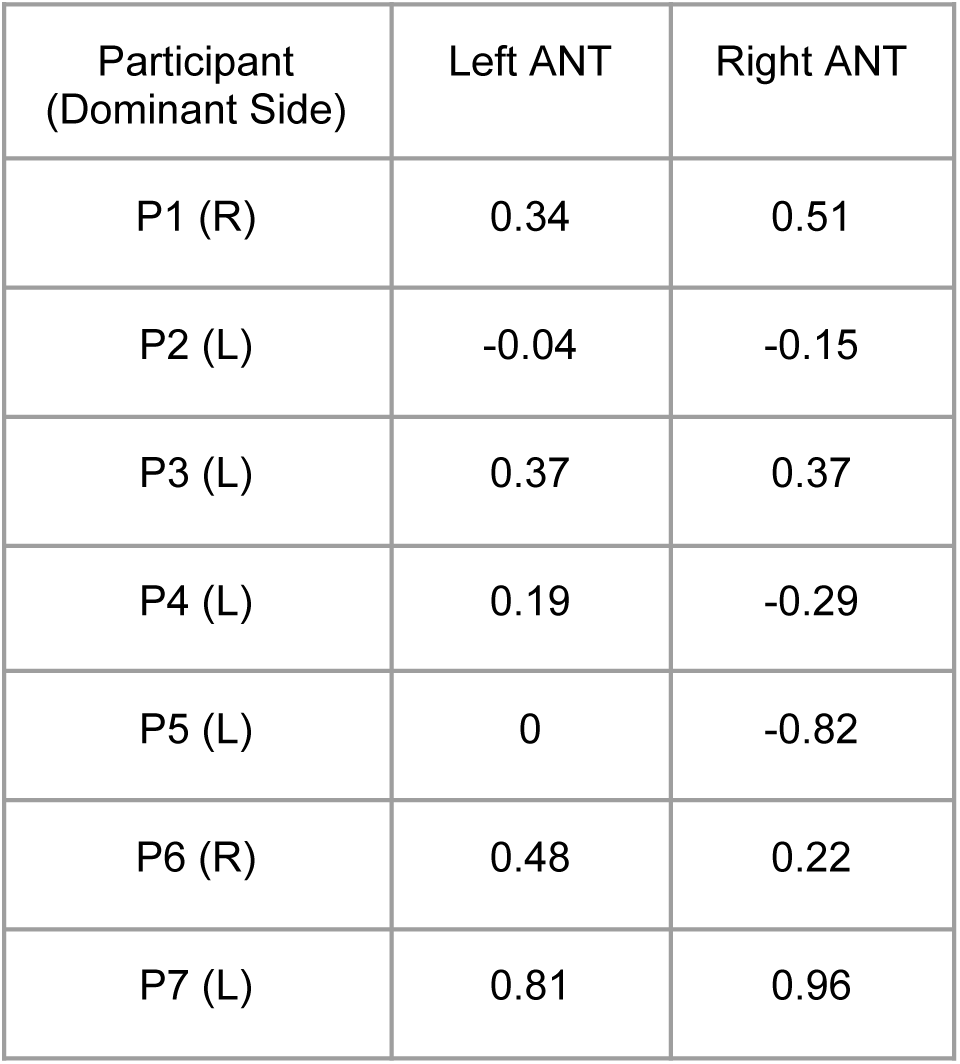
Circadian Amplitude and Seizure Frequency Correlation R-value. Each row contains the 3-point moving average correlation results of LFP in each hemisphere with seizure frequency. Hemisphere with the higher circadian power is indicated in the ‘()’ next to the participant number.

**Supplementary Table 3 -.**
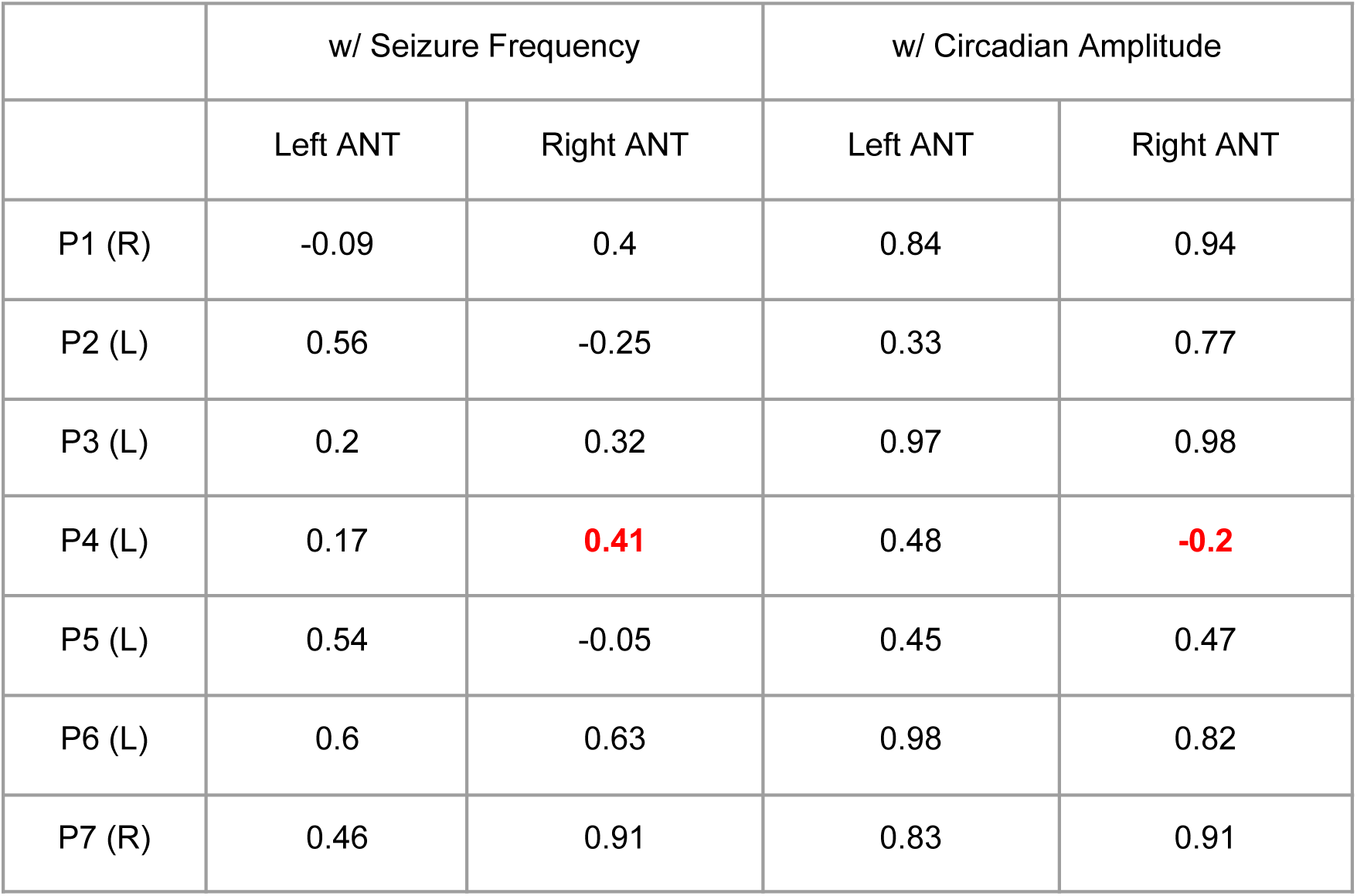
12-hour Cycle Amplitude and Circadian Amplitude/Seizure Frequency Correlation R-value. In addition to the circadian cycle, we observed a strong 12-hour cycle in several participants’ recordings. Most participants showed strong correlations between their circadian and 12-hour cycles in ANTs. However, a negative correlation was found in P4’s right hemisphere (red & bold). This indicates that the circadian and 12-hour cycles could be modulated by different underlying mechanisms. A positive correlation between 12-hour cycle power and seizure frequency was found in most participants, consistent with our findings on the circadian cycle power modulation.

|  | w/ Seizure Frequency |  | w/ Circadian Amplitude |  |
| --- | --- | --- | --- | --- |
|  | Left ANT | Right ANT | Left ANT | Right ANT |
| P1 (R) | -0.09 | 0.4 | 0.84 | 0.94 |
| P2 (L) | 0.56 | -0.25 | 0.33 | 0.77 |
| P3 (L) | 0.2 | 0.32 | 0.97 | 0.98 |
| P4 (L) | 0.17 | **0.41** | 0.48 | **-0.2** |
| P5 (L) | 0.54 | -0.05 | 0.45 | 0.47 |
| P6 (L) | 0.6 | 0.63 | 0.98 | 0.82 |
| P7 (R) | 0.46 | 0.91 | 0.83 | 0.91 |

**Supplementary Table 4 -.** Circadian Cycle Modulation and Stimulation Parameters. Results of modeling monthly circadian cycle power and stimulation parameters using all participants’ data. Significant associations between time, pulse width, and circadian power in the dominant hemisphere, with the stronger modulation over time, were found (p<0.05). Circadian power was z-scored within participants. A 0.01 estimate of pulse width indicates that a 1μs reduction in pulse width is associated with a 0.01 standard deviation reduction in circadian power.

| DominantHemisphereCircadianAmplitude ~ Time + Amps + PW + Frequency + (1\|Participant) |  |  |  |  |  |  |  |
| --- | --- | --- | --- | --- | --- | --- | --- |
| Variable | Estimate | SE | tStat | DF | pValue | Lower CI | Upper CI |
| Intercept | 0.14 | 0.81 | 0.17 | 92 | 0.86 | -1.47 | -0.04 |
| **Time (Month)** | -0.07 | 0.01 | -5.76 | 92 | **<.001** | -0.9 | -0.04 |
| Amps (mA) | 1e-4 | 0.06 | 0.002 | 92 | 1 | -0.11 | 0.11 |
| **PW (μs)** | 0.01 | 0.003 | 2.46 | 92 | **0.02** | 0.001 | 0.01 |
| Frequency (Hz) | -0.001 | 0.005 | -0.29 | 92 | 0.77 | -0.01 | 0.008 |

**Supplementary Table 5 -.** Participant Deep Brain Stimulation Parameters. The dominant hemisphere was selected based on which hemisphere exhibited a stronger circadian cycle across time. Circadian amplitude was z-scored.

| Participant | Time (Month) | CircadianAmp Dominant | Seizure Frequency | Amps | PW | Freq |
| --- | --- | --- | --- | --- | --- | --- |
| 1 | 1 | 0.55 | 5.00 | 1.5 | 120 | 145 |
|  | 2 | 0.25 | 6.33 | 1.5 | 120 | 145 |
|  | 3 | 0.34 | 5.00 | 3 | 90 | 145 |
|  | 4 | 0.83 | 4.33 | 3 | 90 | 145 |
|  | 5 | 1.03 | 1.33 | 3 | 90 | 145 |
|  | 6 | 0.78 | 3.33 | 3 | 90 | 145 |
|  | 7 | -0.14 | 2.33 | 3 | 90 | 145 |
|  | 8 | -0.49 | 3.00 | 4.3 | 50 | 125 |
|  | 9 | -0.25 | 1.00 | 4.3 | 50 | 125 |
|  | 10 | -0.09 | 1.00 | 4.3 | 50 | 125 |
|  | 11 | -0.40 | 1.00 | 3 | 90 | 145 |
|  | 12 | -1.09 | 0.67 | 4.3 | 50 | 125 |
|  | 13 | -1.44 | 1.00 | 4.3 | 50 | 125 |
| **2** | 1 | 0.37 | 4.00 | 4 | 90 | 145 |
|  | 2 | 0.05 | 3.33 | 4 | 90 | 145 |
|  | 3 | 0.02 | 4.00 | 5 | 60 | 125 |
|  | 4 | 0.03 | 5.67 | 5 | 60 | 125 |
|  | 5 | 0.33 | 6.67 | 4 | 90 | 145 |
|  | 6 | 1.07 | 7.34 | 4 | 90 | 145 |
|  | 7 | 1.62 | 7.34 | 4 | 90 | 145 |
|  | 8 | 1.57 | 8.00 | 4 | 90 | 145 |
|  | 9 | 0.60 | 7.67 | 4 | 90 | 145 |
|  | 10 | -0.48 | 6.67 | 4 | 90 | 145 |
|  | 11 | -0.78 | 7.34 | 4 | 90 | 145 |
|  | 12 | -0.81 | 7.67 | 4 | 90 | 145 |
|  | 13 | -0.64 | 7.00 | 4 | 90 | 145 |
|  | 14 | -0.48 | 7.34 | 4 | 90 | 145 |
|  | 15 | -0.23 | 8.00 | 4 | 90 | 145 |
|  | 16 | -0.38 | 9.34 | 4 | 90 | 145 |
|  | 17 | -0.69 | 8.76 | 4 | 90 | 145 |
|  | 18 | -0.96 | 8.64 | 4 | 90 | 145 |
| **3** | 1 | 1.92 | 2.50 | 2 | 90 | 145 |
|  | 2 | 1.39 | 1.67 | 3 | 90 | 145 |
|  | 3 | 1.28 | 2.00 | 3 | 90 | 145 |
|  | 4 | 0.67 | 1.00 | 1.7 | 140 | 165 |
|  | 5 | 0.97 | 1.33 | 1.7 | 140 | 165 |
|  | 6 | 0.19 | 0.67 | 1.7 | 140 | 165 |
|  | 7 | 0.01 | 0.33 | 1.7 | 140 | 165 |
|  | 8 | -0.27 | 1.00 | 1.7 | 140 | 165 |
|  | 9 | 0.00 | 2.33 | 1.7 | 140 | 165 |
|  | 10 | 0.14 | 3.00 | 1.7 | 140 | 165 |
|  | 11 | 0.00 | 2.33 | 2.9 | 60 | 165 |
|  | 12 | -0.02 | 1.67 | 2.9 | 60 | 165 |
|  | 13 | -0.35 | 1.33 | 2.9 | 60 | 165 |
|  | 14 | -0.68 | 1.00 | 2.9 | 60 | 165 |
|  | 15 | -0.98 | 0.33 | 2.9 | 60 | 165 |
|  | 16 | -1.07 | 0.00 | 2.9 | 60 | 165 |
|  | 17 | -1.18 | 0.00 | 2.9 | 60 | 165 |
|  | 18 | -1.11 | 0.67 | 2.9 | 60 | 165 |
|  | 19 | -0.67 | 1.38 | 2.9 | 60 | 165 |
|  | 20 | -0.35 | 2.07 | 2.9 | 60 | 165 |
| 4 | 1 | 0.61 | 3.50 | 2.7 | 90 | 125 |
|  | 2 | 0.26 | 3.67 | 2.7 | 90 | 125 |
|  | 3 | 0.09 | 2.67 | 2.7 | 90 | 125 |
|  | 4 | 1.33 | 2.67 | 2.7 | 90 | 125 |
|  | 5 | 1.45 | 2.67 | 2.7 | 90 | 125 |
|  | 6 | 1.16 | 3.67 | 2.5 | 90 | 145 |
|  | 7 | 0.02 | 3.67 | 2.5 | 90 | 145 |
|  | 8 | -0.09 | 3.33 | 2.7 | 90 | 125 |
|  | 9 | -0.28 | 3.33 | 2.7 | 90 | 125 |
|  | 10 | -0.45 | 3.00 | 2.7 | 90 | 125 |
|  | 11 | -0.46 | 2.67 | 2.7 | 90 | 125 |
|  | 12 | -0.47 | 2.33 | 2.7 | 90 | 125 |
|  | 13 | -0.47 | 3.00 | 3.3 | 60 | 125 |
|  | 14 | -0.48 | 3.67 | 3.3 | 60 | 125 |
|  | 15 | -0.49 | 4.67 | 2.7 | 90 | 125 |
|  | 16 | -0.50 | 5.00 | 2.7 | 90 | 125 |
|  | 17 | -0.51 | 4.00 | 2.7 | 90 | 125 |
|  | 18 | -0.51 | 2.00 | 2.7 | 90 | 125 |
|  | 19 | -0.51 | 0.50 | 2.7 | 90 | 125 |
| 5 | 1 | -0.53 | 66.52 | 4.5 | 90 | 145 |
|  | 2 | -0.51 | 62.35 | 5.9 | 120 | 125 |
|  | 3 | -0.22 | 60.35 | 4.5 | 90 | 145 |
|  | 4 | 0.89 | 44.34 | 5.9 | 120 | 125 |
|  | 5 | 1.47 | 36.34 | 5.9 | 120 | 125 |
|  | 6 | 1.64 | 50.01 | 4.2 | 120 | 125 |
|  | 7 | 0.42 | 46.01 | 5.9 | 60 | 125 |
|  | 8 | -0.24 | 45.34 | 5.9 | 60 | 125 |
|  | 9 | -0.80 | 32.34 | 5.9 | 60 | 125 |
|  | 10 | -0.80 | 32.01 | 5.9 | 60 | 125 |
|  | 11 | -0.70 | 31.01 | 4.2 | 120 | 125 |
|  | 12 | -0.14 | 25.67 | 4.2 | 120 | 125 |
|  | 13 | 0.11 | 23.01 | 4.2 | 120 | 125 |
|  | 14 | 0.10 | 20.67 | 4.8 | 90 | 125 |
|  | 15 | -0.29 | 19.00 | 4.8 | 90 | 125 |
|  | 16 | -0.41 | 16.00 | 4.2 | 120 | 125 |
| **6** | 1 | -0.65 | 15.00 | 4 | 90 | 145 |
|  | 2 | -0.39 | 13.67 | 4 | 90 | 145 |
|  | 3 | -0.34 | 14.34 | 4 | 90 | 145 |
|  | 4 | 0.43 | 12.77 | 4 | 90 | 145 |
|  | 5 | 0.58 | 13.65 | 4 | 90 | 145 |
| **7** | 1 | -0.27 | 5.00 | 1 | 90 | 145 |
|  | 2 | -0.52 | 3.33 | 1 | 90 | 145 |
|  | 3 | -0.42 | 2.67 | 1 | 90 | 145 |
|  | 4 | -0.46 | 1.33 | 1 | 90 | 145 |
|  | 5 | 0.52 | 13.35 | 1 | 90 | 145 |
|  | 6 | 0.87 | 20.02 | 1 | 90 | 145 |

